# Is Footedness Driving Right Lower Extremity Arthroplasty Becoming Commoner Than Left Lower Extremity Arthroplasty?

**DOI:** 10.1101/2021.11.22.21266672

**Authors:** Deepak Gupta

**Author notes:** **Corresponding Author:** Dr Deepak Gupta, Clinical Assistant Professor, Anesthesiology, Wayne State University/Detroit Medical Center, Box No 162, 3990 John R, Detroit, MI 48201, United States.

## Abstract

The question arises whether footedness transforms into asymmetrical incidence of wear and tear within lower extremity joints. After obtaining institutional review board approval for exempt research, the author manually counted the number of patients who underwent right or left or bilateral, hip or knee, primary or revision arthroplasty over a five-year period (2016-2020) at a university-affiliated hospital in the United States. Overall, right lower extremity arthroplasty was significantly commoner than left lower extremity arthroplasty (P=0.002). Individually, only right primary hip arthroplasty (P=0.033) and right revision knee arthroplasty (P=0.041) were significantly commoner procedures than their left counterparts. These results should set up the stage for future investigations into footedness retrospectively and prospectively to rule out if commoner right footedness in itself is driving this asymmetrical incidence of lower extremity arthroplasty or whether automatic transmission vehicular driving is independently contributing to this asymmetrical incidence.

## Introduction

Unlike commonly discussed handedness, footedness is rarely discussed in scientific literature and common media [1-3]. The question arises whether footedness transforms into asymmetrical incidence of wear and tear within lower extremity joints. If that is true, thence the question arises whether left lower extremity only idling and right lower extremity only moving while driving automatic transmission vehicles aggravate such asymmetrical incidence of wear and tear within lower extremity joints [4]. Interestingly, the idea of overuse related wear and tear of joints has been challenged by the idea of disuse related weakening of muscles and bones supporting joints [5-6]. Overall, a simple first step for this inquisition is whether lower extremity arthroplasty is significantly commoner on one side.

## Methods

After obtaining institutional review board approval for exempt research, the author manually counted the number of patients who underwent right or left or bilateral, hip or knee, primary or revision arthroplasty over a five-year period (2016-2020) at a university-affiliated hospital in the United States. The manual counting was done on the onscreen “Case Selection” section in Cerner PowerChart. Only completed cases were counted with case completion indicated by presence of documented surgery stop dates/times on the onscreen “Case Selection” section in Cerner PowerChart.

## Results

Out of 31247 orthopedic and non-orthopedic cases boarded in the onscreen “Case Selection” section in Cerner PowerChart over the five-year period (2016-2020), 4690 completed lower extremity arthroplasty cases were manually counted by the author. Their yearly distribution as depending on side, site and type is depicted in Figure 1. After adding the number of bilateral primary knee arthroplasty (n=34) into both right primary knee arthroplasty as well as left primary knee arthroplasty, the cumulative numbers of lower extremity arthroplasty for the five-year period were tabulated as depicted in Table 1. Overall, right lower extremity arthroplasty was significantly commoner than left lower extremity arthroplasty (P=0.002) per 2×1 Chi-Square analysis. Individually, only right primary hip arthroplasty (P=0.033) and right revision knee arthroplasty (P=0.041) were significantly commoner procedures than their left counterparts per 2×1 Chi-Square analysis.

**Table 1:** Comparison Among Lower Extremity Arthroplasty: Right-Sided Vs. Left-Sided

| **Arthroplasty** | **Cumulative Numbers for 5-Years**<br>**(2016-2020)** | **P-Value** |
| --- | --- | --- |
| Right HIP Primary | 820 | **0.033** |
| Left HIP Primary | 736 |  |
| Right HIP Revision | 125 | 0.237 |
| Left HIP Revision | 107 |  |
| Right KNEE Primary | 1322 | 0.15 |
| Left KNEE Primary | 1249 |  |
| Right KNEE Revision | 202 | **0.041** |
| Left KNEE Revision | 163 |  |
| RIGHT LOWER EXTREMITY | 2469 | **0.002** |
| LEFT LOWER EXTREMITY | 2255 |  |

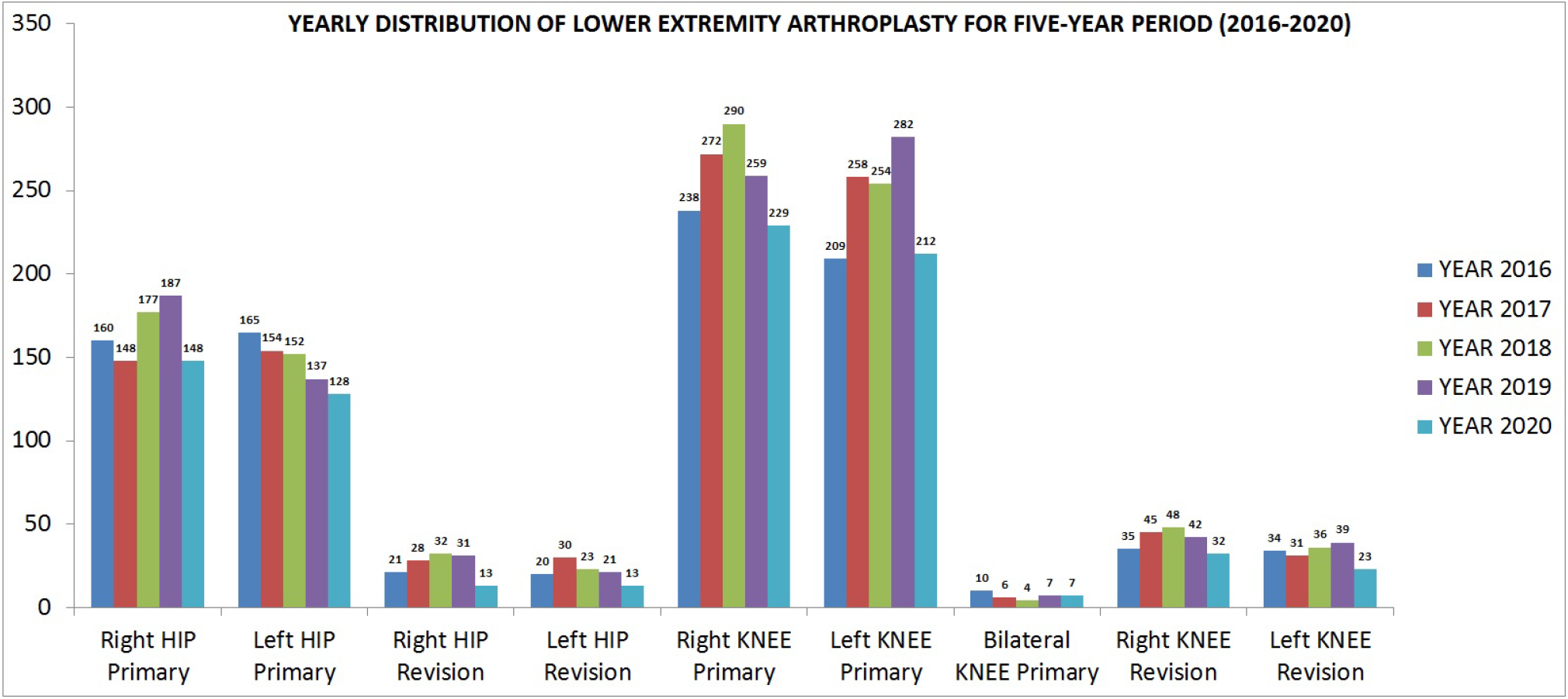

## Discussion

The author’s null hypothesis about symmetrical incidence of arthroplasty in lower extremity was rejected by commoner incidence of right lower extremity arthroplasty. These results should set up the stage for future investigations into footedness retrospectively and prospectively to rule out if commoner right footedness in itself is driving this asymmetrical incidence of lower extremity arthroplasty or whether automatic transmission vehicles’ driving is independently contributing to this asymmetrical incidence.

There were some study limitations. The multi-thousand patients’ data was only superficially and manually counted to decipher just the cumulative numbers of lower extremity arthroplasty over the five-year period. Comprehensive retrospective chart review of 4690 cases could not be planned by the lone author-researcher. Future retrospective and prospective studies by larger research teams may be able to address this logistical limitation.

## Supporting information

IRB exemption letter

## Data Availability

All data produced in the present work are contained in the manuscript.

## Data Availability

All data produced in the present work are contained in the manuscript.

## Acknowledgement

The author is indebted to Shushovan Chakrabortty, MD, PhD, Department of Anesthesiology and Pain Medicine, Wayne State University, Detroit, Michigan, United States, for introducing the thought of footedness into my scientific quest and its exploration.

## Notes

**Financial Support:** None

**Conflicts of Interests:** None

### Competing Interest Statement

The authors have declared no competing interest.

### Funding Statement

This study did not receive any funding

### Author Declarations

Wayne State University: IRB Administration Office gave ethical approval as concurrence of exemption for this work.

