## Supplementary material for "Is Footedness Driving Right Lower Extremity Arthroplasty Becoming Commoner Than Left Lower Extremity Arthroplasty?": IRB exemption letter

IRB Administration Office
87 East Canfield, Second Floor
Detroit, MI 48201
www.irb.wayne.edu

**CONCERRENCE**

### CONCURRENCE OF EXEMPTION
#### IRB-21-08-3904-MP2 Expedited/Exempt-EXEMPT

| | |
|---|---|
| **DATE:** | October 11, 2021 |
| **TO:** | Gupta, Deepak, Anesthesiology |
| | Zestos, Maria, Anesthesiology |
| **FROM:** | Smitherman, Lynn, Associate Professor - Clinical, MP2 Expedited/Exempt |
| **PROTOCOL TITLE:** | Is Right Lower Extremity Arthroplasty More Common Than Left Lower Extremity Arthroplasty At Our Institute? |
| **FUNDING SOURCE:** | None |
| **PROTOCOL NUMBER:** | IRB-21-08-3904 |
| | Approval Date: October 07, 2021 |

The above-referenced protocol has been reviewed and found to qualify for Exemption according to category 4

[

**Note to PI:** This IRB approval does not replace administrative or department/college/division approvals that may be required. Before initiating research activities contact the Associate/Vice Dean for Research in your school or college for established parameters for site access to the facility where the study will be conducted.

**NOTE TO PRINCIPAL INVESTIGATORS**: Due to the COVID-19 health crisis, the resumption of human participant research is occurring in measured phases which incorporate institutional, state, and federal regulations and best practices.

Currently the following research activities are ongoing:

(I) Human participant research that can maintain remote study interventions/visits as per IRB approval.

(II) In-person study visit(s) that coincide with a standard of care visit(s). Note, study visits that have been modified to virtual/phone/remote formats can continue to conduct those visits as per IRB approval.

(III) In-person research that can provide a potential direct benefit to the participant.

(IV) **Effective June 1, 2021:** In-person research that has no potential for direct benefit conducted at Wayne State University sites and/or established health care facilities.

(V) **Effective July 1, 2021:** In-person research without potential for direct benefit to participants conducted at non-affiliated WSU sites will resume. The study's mitigation plan must include the non-affiliated study sites' approval/letter of support to conduct in-person research activities. This approval/letter must address approval/support to resume research activities at their site effective July 1, 2021.

In-person research activities require additional precautions to protect both the participant and the research staff. The Principal Investigator should review the IRB 's Appendix N: Resumption of In-Person Clinical Research Form for instructions. For more information regarding Appendix N and IRB resumption of research requirements visit: research.wayne.edu/irb/coronavirus.

WAYNE STATE UNIVERSITY

IRB Administration Office
87 East Canfield, Second Floor
Detroit, MI 48201
www.irb.wayne.edu

When clinical research is conducted in a standard medical care/hospital setting, please follow that site ’s COVID-19 precautionary standard operating procedures Appendix N is not required.

For research conducted at a WSU research facility, refer to the university guidance. Information on restarting WSU research operations can be found at: research.wayne.edu/coronavirus/restart guidance.

For more information regarding IRB submission requirements and instructions visit the IRB Forms and Submissions Requirements website: research.wayne.edu/irb/forms-requirements-categories. For more information about the phased resumption of human participant research visit: research.wayne.edu/irb/coronavirus. .

If you have questions please contact the IRB Administration Office, or telephone: 313-577-1628.
The following attachments and consent/assent documents have been reviewed and approved by the IRB.

Notes:
NOTE TO PI: This project has been given a Status Check-In Date. The Status Check-In Date is 10/06/2023. The Minimal Risk Status Update Form should be used to provide a status report to the IRB. Please submit the status update at least 6 weeks before this date. The Minimal Risk Status Update Form is available on the IRB's Forms and Submissions website (www.irb.wayne.edu). The Minimal Risk Status Update should be submitted as an expedited amendment via eProtocol with the Minimal Risk Status Update Form. Include the Minimal Risk Status Update Form as an Attachment using the label: Minimal Risk Status Update.

Protocol/Proposal/Dissertation (dated 08/2021)

The following data collection materials have been reviewed and approved and does not require a WSU IRB stamp for use. These documents are approved and noted in the IRB file (1): Data Collection Sheet.

A waiver of consent has been granted according to 45CFR46.116(d). This waiver satisfies: 1) risk is no more than minimal, 2) the waiver does not adversely affect the rights and welfare of research participants, 3) the research could not be practicably carried out without the waiver, and 4) the participants will not be given information.

A waiver of HIPAA Authorization has been granted in accordance with the Privacy Rule and Justification provided by the Principal Investigator in the HIPAA Summary Form. This waiver satisfies: 1) the use or closure of PHI involves no more than minimal risk to the privacy of individuals, 2) the research could not be practicably conducted without the waiver, 3) the research could not be practicably conducted without access and use of the PHI, 4) adequate steps taken to protect identifiers from improper use of disclosure and 5) adequate plan for destroying identifiers or links.

Effective September 1, 2021: For research conducted at WSU Campus sites: Participants must complete the WSU Campus Guest Screener. The WSU campus vaccine mandate is not required for research participants who are visiting campus to participate in a research study. However, mitigation plans as indicated for Appendix N must be followed. The research participant must be contacted before the study visit to inform them of the WSU campus screening and safety precautions.

WAYNE STATE UNIVERSITY

IRB Administration Office
87 East Canfield, Second Floor
Detroit, MI 48201
www.irb.wayne.edu

* Exempt protocols do not require annual review by the IRB, however you may have been granted a Status Check-In Date. Projects granted a Status Check-In date must submit a Minimal Risk Status Update Report at least 6 weeks before the check-In date. If research activities are complete a Final Report/Closure must be submitted by the Status Check-In date.

* All changes or amendments to the above-referenced protocol require review and approval by the IRB BEFORE implementation.

* Adverse Reactions/Unanticipated Problems AR/UP must be submitted on the appropriate form within the time frame specified in the IRB. In the event of an unanticipated problem use the Unanticipated Problem Report Form located on the IRB's Forms and Submissions Requirements website.

Note: Studies conducted at DMC sites or DMC medical record used for affiliate review Authorized DMC personnel have been added to this submission under Personnel Information "Other".
Administration Office Policy www.irb.wayne.edu/policies-human-research

NOTE: Upon notification of an impending regulatory site visit, hold notification, and/or external audit the IRB Administration Office must be contacted immediately. Also Notify the IRB of any changes to the funding status of the above-referenced protocol.

**Attachments**

- Protocol REVISION 1
- DMC 19833 DMC CRO Review form
- Research Review Authorization
- DATA WORKSHEET
- Protocol
- Concise CV Deepak Gupta

---

| | |
|---|---|
| **Review Type:** | EXEMPT |
| **IRB Number:** | MP2 Expedited/Exempt Review |
